# Age-related changes, influencing factors and crosstalk between vaginal and gut microbiota: a cross-sectional comparative study of pre- and postmenopausal women

**DOI:** 10.1101/2021.12.22.21268221

**Authors:** Remi Yoshikata, Michiko Yamaguchi, Yuri Mase, Ayano Tatuzuki, Khin Z. Myint, Hiroaki Ohta

**Author notes:** Corresponding author and to whom reprint request should be addressed: Khin Zay Yar Myint, Tokyo Midtown Medical Center, 9-7-1, Akasaka, Minato-ku, Tokyo, 107-6206.

## Abstract

**Objective:** The ideal vaginal environment is maintained by Lactobacillus species, which keep the vagina clean and free of infections, including sexually transmitted diseases and human papilloma virus infection. Other reported health benefits of *Lactobacillus* include a favorable impact on fertility and immunity, leading to a reduced risk of gynecological malignancies. Age-related decline in estrogen affects the population of *Lactobacillus*, leading to dominance of pathogenic flora and increased diversity in vaginal microbiota. In this study, we compared the differences between the vaginal microbiota of premenopausal and postmenopausal women. In addition, we examined the relationships between vaginal and gut microbiota, as well as their relationships to sex hormones and equol producing ability.

**Methods:** This was a cross-sectional study of 35 premenopausal and 35 postmenopausal women, ranging from 27 to 76 years of age. We compared parameters such as the composition of the gut and vaginal microbiota, vaginal pH, sex hormones in the blood (estradiol and follicular stimulating hormone), and urinary equol concentration.

**Results:** In the vaginal microbiota of premenopausal women, *Lactobacillus* species constituted approximately 71.98%, and pathogenic flora constituted approximately 16.87%. They were 10.08% and 26.78%, respectively, in the vaginal microbiota of postmenopausal women. Therefore, the proportion of *Lactobacillus* was significantly low, whereas microbial diversity and vaginal pH were significantly high (p<0.0001) in postmenopausal women. The compositions of the vaginal microbiota were significantly different in pre- and postmenopausal women. However, such differences were not noticeable in the gut microbiota. Urinary equol production had no significant correlation with vaginal microbiota, although it had significant relationships with gut microbiota in postmenopausal women. There were significant correlations among bacterial species in the gut and vaginal microbiota, especially in postmenopausal women. In both groups, the proportions of vaginal *Lactobacillus* were inversely correlated with vaginal microbial diversity and vaginal pH.

**Conclusion:** Postmenopausal women had significantly low *Lactobacillus* and high pathogenic flora in their vaginal flora, whereas such age-related differences were not identified in gut microbiota. There were significant correlations among the bacterial species inhabiting the gut and vaginal microbiota, especially in postmenopausal women, indicating potential crosstalk between each other.

## Introduction

*Lactobacillus* species are gram-positive bacteria belonging to the phylum Bifidobacterium. A wide variety of *Lactobacillus* species exist in the world, ranging from those in fermented food as well as in humans and animals. In humans, *Lactobacillus* species constitute the normal flora of the mouth, gastrointestinal tract and female vagina. Vaginal *Lactobacillus* species, through their natural protective actions, play a major role in maintaining the ideal vaginal environment and vaginal health.

The *Lactobacillus* population in the vagina is influenced by blood estrogen levels. Estrogen stimulates the production of glycogen in the vaginal epithelium, and *Lactobacillus* is produced by the action of amylase and vaginal microbiota on shedding vaginal epithelium. The functions of *Lactobacillus* are killing and dissolving the membranes of pathogenic bacteria by creating an acidic vaginal environment and stimulating immunity similar to an antibiotic. The synergistic effect of estrogen with *Lactobacillus* species creates the first line of defense mechanism against the proliferation of pathogens. The eubiotic effects of estrogen and *Lactobacillus* species in the vaginal ecosystem were summarized by Amabebe and Anumba in 2018 (Supplementary data 1). ^1^

There are five community types or community state types (CSTs) of *Lactobacillus* in the vagina.^2^ CST I, CST II, CST III and CST V are dominated by L. crispatus, L. gasseri, L. iners, and L. jensenii, respectively. The ranking of *Lactobacillus* was in the order of CST I, II, V, and III; the better the rank was, the lower the risk of sexually transmitted infections and viral infections, such as HIV, HSV, HPV, gonorrhea, chlamydia, and trichomonas. The remaining type is the CST IV or diversity type, which is dominated by highly diverse anaerobes with low proportions of typical Lactobacillus species. CST IV can be further classified into CST IV-A and CST IV-B, depending on the composition of bacteria. ^3^ Several studies have reported that CST IV is associated with bacterial vaginosis, sexually transmitted infection, human papilloma virus infection, reduced fertility, and a negative effect on immunomodulation leading to gynecological cancers. ^4-6^ Therefore, diversity of the vaginal microbiota is associated with an increased risk for various gynecological problems.

CST I is thought to be the most prevalent species, which maintains vaginal pH no more than pH 4.4. CST IV is mostly made up of anerobic bacteria, which maintain a vaginal pH greater than 5.3.^2^ The prevalence of these subtypes is different across different ethnic groups or geographical locations. CST I, II, III and V make up 73% and 90% of White and 80% of Asian populations, and CST IV makes up approximately 27% and 40% of Black and 38% of Hispanic populations.^2^ Furthermore, CST prevalence can be temporary; it can vary in the same individual over time, depending on the blood level of estrogen. Therefore, the reduction in intrinsic estrogen due to aging processes can increase microbial diversity due to an imbalance between *Lactobacillus* and pathogenic flora.

In this study, we aimed to investigate the effects of aging on the composition and diversity of the gut and vaginal microbiota using next-generation sequencing technology. In addition, we also examined their relationships to other parameters, such as female sex hormones and equol producing ability.

## Methods

### Study population

This is the baseline cross-sectional study of an interventional study comprising 70 healthy Japanese women who visited the Hamasite Clinic in Tokyo Prefecture from May to July 2021. Half of them were premenopausal women, and half were postmenopausal women. Menopause was defined as no menstruation at least 12 months since the last menstrual period, follicular stimulating hormone level of 25 pg/ml and above and estradiol level less than 29 pg/ml. Exclusion criteria included those currently having or being treated for genitourinary symptoms such as vaginitis and cystitis and those around menopause with unstable ovarian function. The Institutional Review Board of Medical Corporation Shinkokai approved the entire research protocol. All participants provided written informed consent for participating in the study. This study was registered with the University Hospital Medical Information Network (UMIN) Clinical Trial Registry (trial registration number: UMIN000043944).

### Measurements

Female sex hormones such as estradiol and follicular stimulating hormone levels were measured from the blood specimens of all participants. As there can be physiological fluctuations in women of reproductive age, we collected blood during the ovulation phase in premenopausal women. Gynecologists, who are involved as coinvestigators of this study, took the required samples and pH measurements from the vagina. Vaginal pH was determined by using pH test strips (pH⍰Fix 3.6–6.1, Macherey-Nagel, Düren, Germany). The measurement of urinary equol was conducted as described by Yoshikata et al. ^7^ Women with a urinary equol level higher than 1.0 μM were defined as equol producers. ^8,9^

### Microbiome tests

For vaginal and fecal specimens, bacterial cell walls were disrupted by the bead-beating method using zirconia beads to extract or isolate bacterial DNA. The extracted DNA was then purified using an automatic extraction device (Maxwell, Promega). Using this extracted DNA as a template, approximately 300 base pairs in the V1 and V2 regions of the 16S rRNA fragments were amplified using a thermal cycler (Veriti, Thermo Fisher Scientific, MA, USA). The amplified products were then purified and quantified using a real-time PCR device (StepOne™, Thermo Fisher Scientific, MA, USA). A library was prepared using the quantified PCR product, and emulsion PCR was performed using special beads by the OneTouch™ system (Thermo Fisher Scientific, MA, USA). After purifying the special beads, they were loaded into an electronic chip and sequenced using a next-generation sequencer (PGM, Thermo Fisher Scientific, MA, USA). The phylum, class, order, family, genus, and species levels were identified, and the proportion of each species was calculated from the obtained sequences. The details of the protocol were described in previous studies. ^10, 11^ Fecal microbiome test results were reported as microbial diversity, FB ratio, and distributions of bacterial groups at the phylum, class, order, family, genus, and species levels. Vaginal microbiome tests were reported as microbial diversity and distributions of bacterial groups at the phylum, class, order, family, genus, and species levels.

The alpha diversity scores of the Shannon diversity index were used to describe microbial diversity. The FB ratio refers to the ratio of the phylum Firmicutes to the phylum Bacteroidetes.

### Statistical analyses

The calculations and figure generation were performed using Microsoft Excel (Microsoft Corporation, 2018) and R software (R 4.1.0, R Core Team, 2021). The distribution of normality was assessed with the Kolmogorov–Smirnov test, box plots, and histograms. Categorical variables were expressed as numbers (N) and percentages (%), and continuous variables were expressed as medians. Comparisons between premenopausal and postmenopausal women were performed using the chi-squared test and Fisher’s exact test for proportions and the Mann–Whitney test for continuous variables. The correlations between the composition of gut and vaginal microbiota and other parameters were evaluated by Spearman’s correlation tests. All tests were two-sided, and statistical significance was set to p < 0.05.

## Results

Table 1 shows the characteristics of the participants in our study cohort. The average urinary concentration of equol was higher, but the proportion of equol producers was lower in postmenopausal women, although these findings were not statistically significant. Nevertheless, age-related changes, such as sex hormone levels and vaginal pH, were significantly different between the two groups. *Lactobacillus* species were the most prominent organisms (average ± SD: 72% ± 36.84) in the vaginal microbiome of premenopausal women, whereas their proportions were approximately 10% (± 25.11) in postmenopausal women. In premenopausal women, *L. iners* (CST III) accounted for approximately 35%, followed by *L. crispatus* (CST I, 28.9%), *L. gasseri* (CST II, 5%) and *L. jensen*i (CST V, <1%), while the pathogenic flora accounted for approximately 16.7%. In postmenopausal women, *L. iners* (CST III) made up approximately 4.8%, followed by *L. gasseri* (CST II, 1.6%), *L. crispatus* (CST I, 1.1%), and *L. jenseni* (CST V) at less than 1%, whereas the pathogenic flora was approximately 26.8%. There were significant differences in the *Lactobacillus* composition as well as vaginal microbial diversity (0.77 versus 1.92) between the two groups of women. However, there were no significant differences in the composition and microbial diversity of the gut microbiome in either group.

**Table 1.** Summary of characteristics of the participants in the study cohort.

|  | Premenopausal women<br>(n=35) |  | Postmenopausal women<br>(n=35) |  | p-value |
| --- | --- | --- | --- | --- | --- |
| Age, years | 37.26 | ± 6.09 | 59.86 | ± 4.81 | <.0001 |
| Height, centimeter | 154.26 | ± 27.19 | 156.74 | ± 4.98 | 0.074 |
| Weight, kilograms | 52.17 | ± 12.97 | 55.12 | ± 8.01 | 0.51 |
| Parity, times | 0.34 | ± 0.76 | 1.66 | ± 1.11 | <.0001 |
| Age of menarche, years | 12.03 | ± 2.55 | 12.55 | ± 1.39 |  |
| Age of menopause, years |  |  | 50.80 | ± 4.54 |  |
| Menstrual period, days | 5.71 | ± 1.86 | 5.87 | ± 0.83 |  |
| Blood |  |  |  |  |  |
| FSH | 6.62 | ± 9.41 | 58.89 | ± 20.66 | <.0001 |
| E2 | 168.10 | ± 261.06 | 10.00 | ± 0.00 | <.0001 |
| Urine |  |  |  |  |  |
| Urinary equol concentration, µM | 4.47 | ± 8.71 | 8.63 | ± 15.92 |  |
| Equol producer status |  |  |  |  | 0.9183 |
| Equol non-producer | 19 | (54%) | 21 | (62%) |  |
| Equol producer | 16 | (46%) | 14 | (41%) |  |
| Vagina |  |  |  |  |  |
| Vaginal pH | 4.37 | ± 0.58 | 5.89 | ± 0.45 | <.0001 |
| Vaginal microbiome |  |  |  |  |  |
| Microbial diversity | 0.77 | ± 0.72 | 1.92 | ± 1.01 | <.0001 |
| Total *Lactobacillus*, % | 71.98 | ± 36.84 | 10.08 | ± 25.11 | <.0001 |
| *L. crispatus* (CST I) | 28.86 | ± 41.89 | 1.10 | ± 6.39 | 0.001 |
| *L. gasseri* (CST II) | 5.02 | ± 14.37 | 1.59 | ± 7.66 | 0.033 |
| *L. iners* (CST III) | 35.02 | ± 41.39 | 4.80 | ± 19.43 | 0.00035 |
| *L. jensenii* (CST V) | 0.48 | ± 1.86 | 0.00 | ± 0.02 |  |
| Pathogenic bacteria (CST IV) | 16.87 | ± 27.35 | 26.78 | ± 29.60 | 0.34 |
| Others | 11.14 | ± 21.39 | 63.14 | ± 34.39 | <.0001 |
| Phylum composition |  |  |  |  |  |
| Firmicutes, % | 48.44 | ± 15.54 | 48.15 | ± 14.5 | 0.78 |
| Bacteroides, % | 51.56 | ± 15.54 | 51.85 | ± 14.5 | 0.78 |
| Bifidobacteria, % | 4.69 | ± 5.35 | 3.52 | ± 4.72 | 0.32 |
| Gut microbiome |  |  |  |  |  |
| Microbial diversity | 4.4 | ± 0.48 | 4.41 | ± 0.38 | 0.93 |
| Firmicutes, % | 48.44 | ± 15.54 | 48.15 | ± 14.5 | 0.78 |
| Bacteroides, % | 51.56 | ± 15.54 | 51.85 | ± 14.5 | 0.78 |
| Bifidobacteria, % | 4.69 | ± 5.35 | 3.52 | ± 4.72 | 0.32 |
| Lactobacillus, % | 3.53 | ± 7.76 | 3.1 | ± 4.09 | 0.73 |
| Butyric acid producing bacteria, % | 10.9 | ± 7.51 | 11.52 | ± 10 | 0.83 |
| Equol producing bacteria, % | 1.66 | ± 2.29 | 1.71 | ± 3.22 | 0.77 |

p-value: chi-squared test and Fisher's exact test for proportions and the Mann–Whitney test for continuous variables

### Relationship between vaginal and gut microbiota

They shared the same five phyla: Firmicutes, Bacteroidetes, Proteobacteria, Actinobacteria, and Fusobacteria. There was a significant difference in the composition of phyla between pre- and postmenopausal women in the vaginal microbiota. Firmicutes was reduced significantly (p=0.0035), and Bacteroidetes and Proteobacteria were increased significantly (p=0.022 and p=0.000056, respectively) in postmenopausal women. However, such a difference was not observed with their gut microbiota (Figure 1).

**Figure 1.**
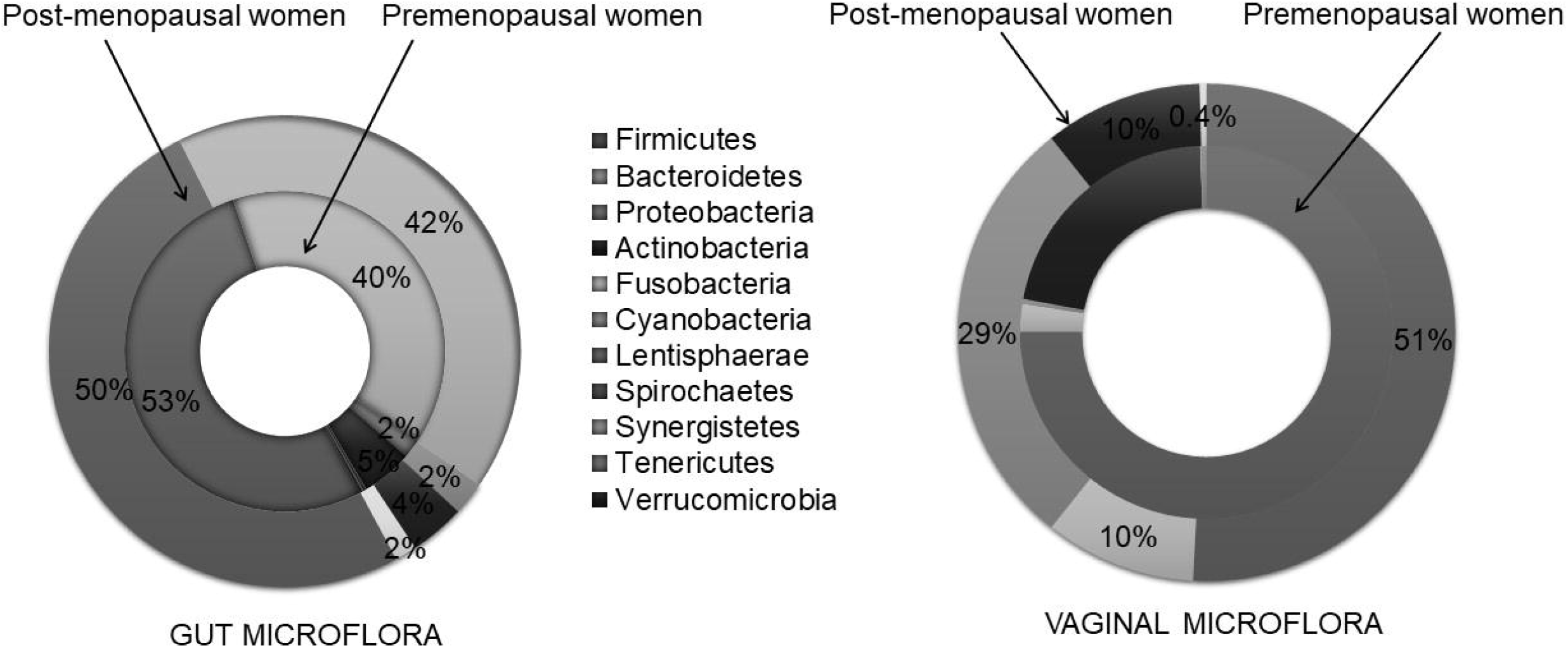
Comparisons of compositions of phylum between pre- and post-menopausal women. There was a significant difference in the composition of phyla between pre- and postmenopausal women in the vaginal microbiota. However, such a difference was not observed with their gut microbiota.

### Comparison of *Lactobacillus* distribution between pre- and postmenopausal women and between vaginal and gut microbiota

Figure 2 shows the distribution of *Lactobacillus* species in each group of women in their vagina and gut. There was no difference in the proportions of *Lactobacillus* species in the gut microbiota of the two groups of women. However, these proportions were significantly different in the vaginal microbiota between the two groups. There were three main *Lactobacillus* species in the vaginal microbiota of premenopausal women. They were

**Figure 2.**
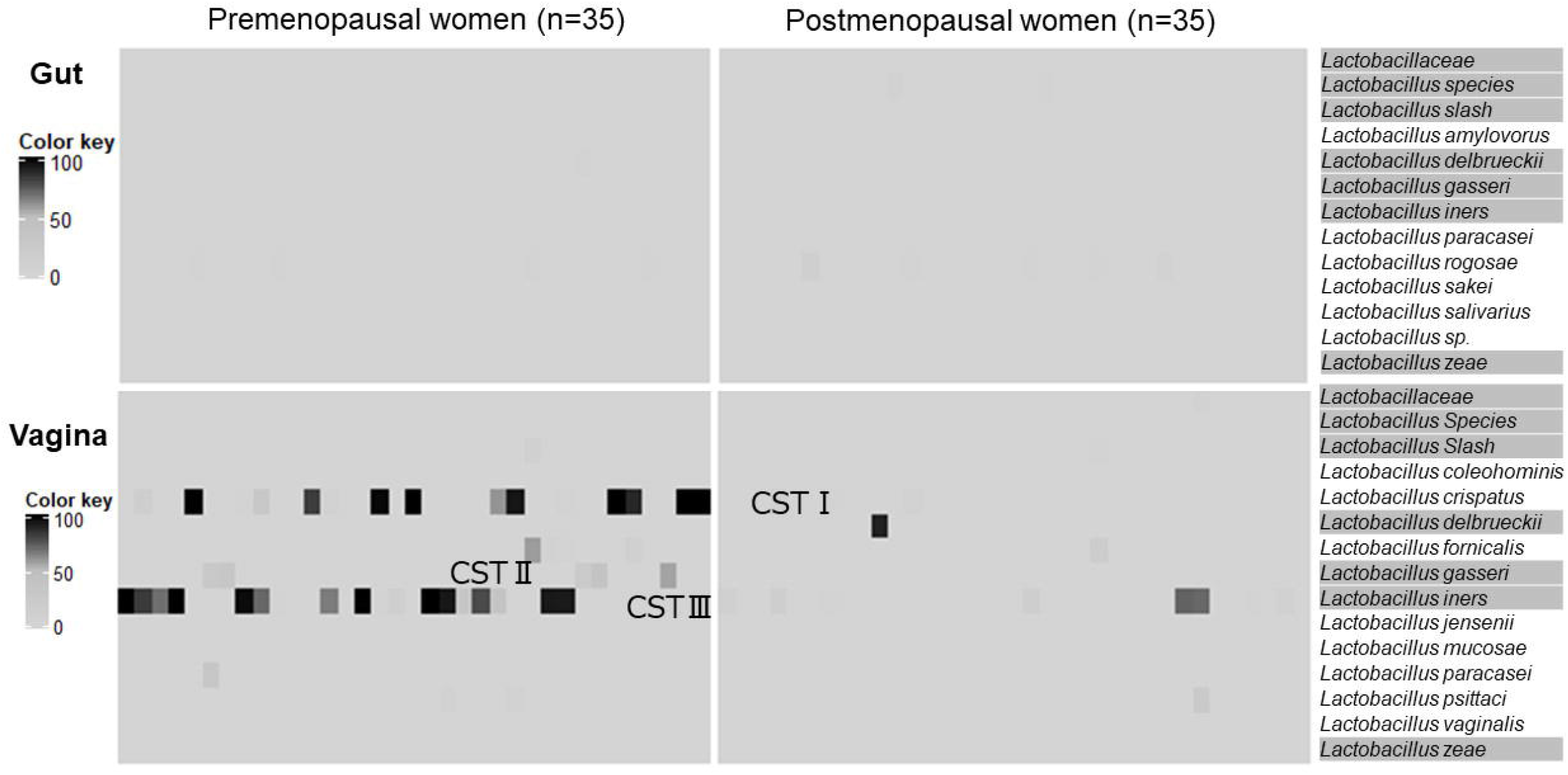
Distribution of *Lactobacillus* species in the gut and vagina. There was no difference in the proportions of *Lactobacillus* species in the gut microbiota of pre- and postmenopausal women. However, these proportions were significantly different in the vaginal microbiota between the two groups. The highlighted bacterial names are the seven common *Lactobacillus* species in the vaginal and gut microbiota in this study cohort.

*L. crispatus* (CST I), *L. iners* (CST III), and *L. gasseri* (CST II). In postmenopausal women, the proportion of *L. crispatus* (CST I) was almost negligible, and *L. iners* (CST III) and *L. gasseri* (CST II) became the main species. The main *Lactobacillus* species in the gut microbiota was *L. rogosae*. There were 7 common *Lactobacillus* species in the vaginal and gut microbiota in this study cohort, and some were specific to the vagina or gut. Among the community state types in this study cohort, *L. iners* (CST III) and *L. gasseri* (CST II) inhabited both the vagina and gut but not *L. crispatus* (CST I).

### Relationships of *Lactobacillus* species between the gut and vagina

Figure 3 shows significant relationships of *Lactobacillus* between the gut and vagina. Some small communities of *Lactobacillus* in the vagina have mutual positive relationships. However, the main community state type of *L. iners* (CST III) in the vagina was not associated with any *Lactobacillus* species in the gut, whereas it was inversely associated with the proportion of *L. crispatus* (CST I) in the vagina. There were positive associations between the proportions of *Lactobacillus paracasei* in the vagina and the gut. *L. gasseri* (CST II) in the vagina was also positively associated with *Lactobacillus* species in the gut. The proportion of *L. rogosae* in the gut was also positively associated with the proportion of *Lactobacillus* species in the vagina. In postmenopausal women, the proportion of the main CST, *L. iners*, in the vagina was positively associated with the proportion of *L. iners* in the gut as well as *Lactobacillaceae* in the vagina. Similarly, the proportion of *Lactobacillus iners* in the gut was positively associated with the proportions of *Lactobacillaceae* and other *Lactobacillus* species in the vagina. Only positive associations were noted among the *Lactobacillus* species in the gut as well as in the vagina.

**Figure 3.**
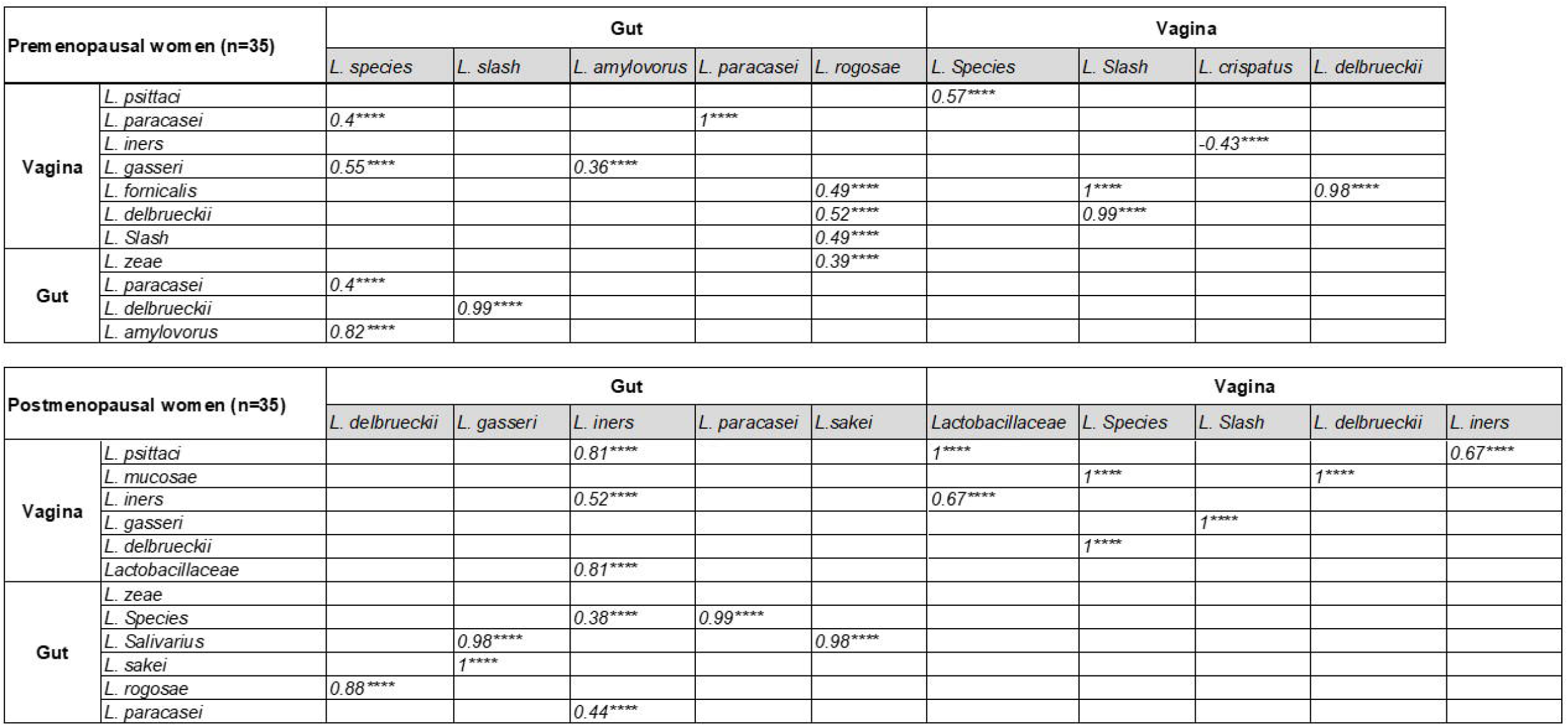
Significant relationships of *Lactobacil/us* species between the gut and vagina. All the significant correlations were described as ****(p<0.0001) in the figure. The main community state type in the premenopausal women was *L. iners* (CST III). It was inversely associated with the proportion of *L. crispatus* (CST I) in the vagina. Only positive associations were noted among the *Lactobacillus* species in the gut as well as in the vagina of the postmenopausal women.

Relationships of common bacteria species in the gut and vagina that are responsible for bacterial vaginosis Figure 4 shows the relationship between the pathogenic flora that are responsible for bacterial vaginosis in the vagina and the gut. In premenopausal women, the proportions of *Bacteroides vulgatus* in the gut were positively correlated with the proportions of most of the pathogenic flora in the vagina. Additionally, the *Prevotella corporis* population in the gut was positively related to other *Prevotella* species in the vagina. Similarly, the vaginal *Prevotella* population had a strong correlation with the gut *Prevotella* species (r>0.9, p<0.0001). There were also strongly positive correlations among the pathogenic bacteria in both the gut and vagina, except for *Bacteroides vulgatus* in the gut, which had a negative association with *Bacteroides dorei*. In postmenopausal women, there were several correlations between the vaginal and gut microbiota that were responsible for bacterial vaginosis. The proportions of *Bacteroides, Prevotella* and *Streptococcus* in the gut were strongly and positively related to those proportions in the vagina. Similar to premenopausal women, the vaginal *Prevotella* population also had a strong relationship with the gut *Prevotella* species (r>0.5, p<0.0001). There were also strongly positive correlations among the pathogenic bacteria in both the gut and vagina, except for *Bacteroides vulgatus* in the gut, which had a negative association with *Bacteroides eggerthii*.

**Figure 4.**
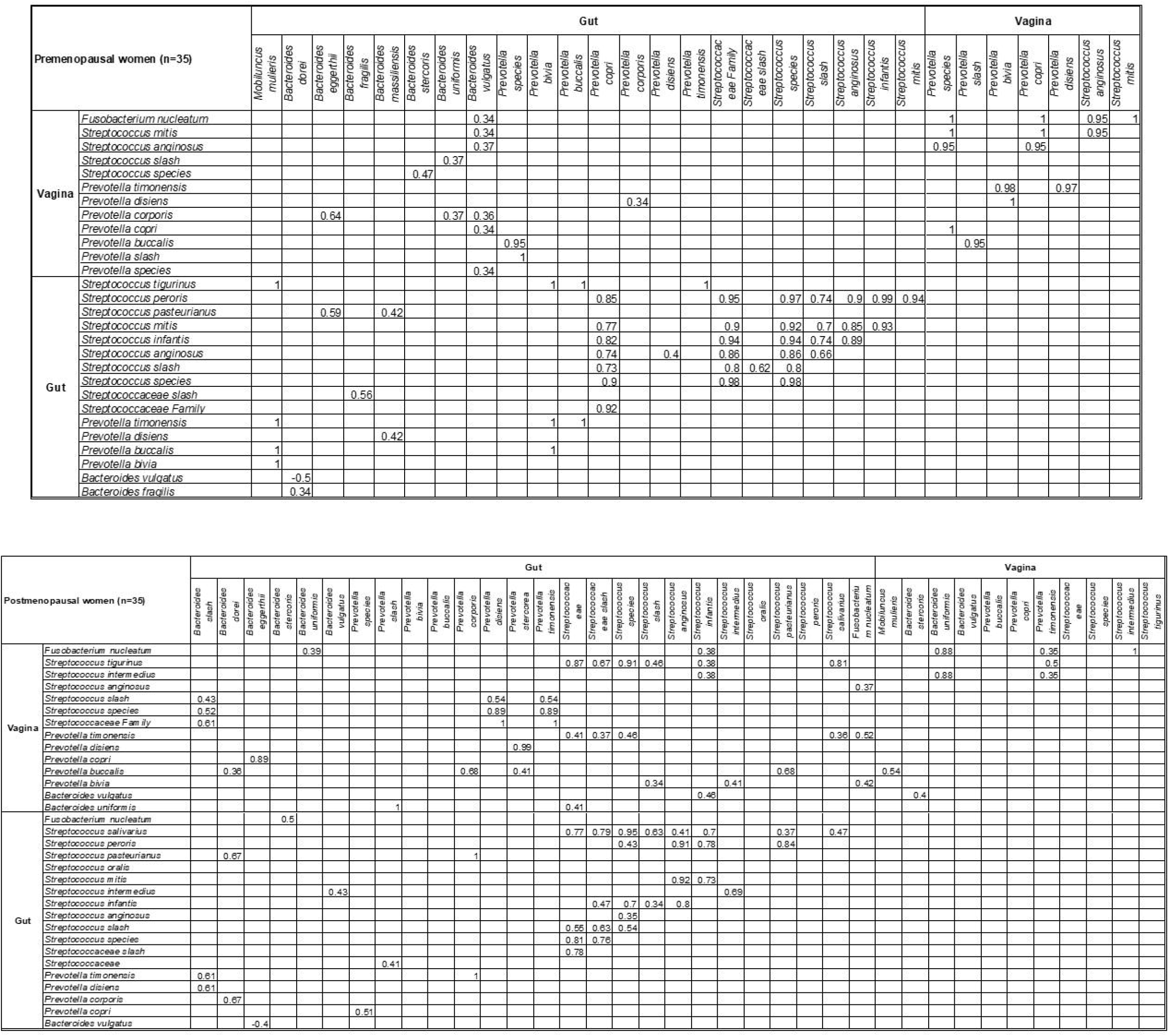
Significant relationships of pathogenic flora between the gut and vagina. Only significant correlations (p<0.05) were described in the figure. There were strongly positive correlations among the pathogenic bacteria in both the gut and vagina, especially in postmenopausal women.

### Relationships of hormonal and other parameters on the gut and vaginal microbiota

Factors affecting the vaginal microbiota are shown in Figures 5A and 5B. In premenopausal women, vaginal *Lactobacillus* proportions were negatively associated with the proportion of pathogenic flora, vaginal pH and microbial diversity. Then, there was a positive correlation between vaginal pH and microbial diversity. In addition, there was a positive correlation between equol-producing bacterial proportion and estrogen, whereas a negative correlation was noted between urinary equol concentration and gut microbial diversity. There was also a negative relationship between Firmicutes and Bacteroides. In postmenopausal women, such as premenopausal women, there was a negative relationship between the proportion of vaginal *Lactobacillus* and vaginal pH, where vaginal pH was positively associated with microbial diversity. In the gut microflora, there was a negative relationship between Firmicutes and Bacteroides, a positive relationship between urinary equol and Firmicutes, and a negative relationship between urinary equol and Bacteroides. However, there was a positive relationship between gut microbial diversity and Firmicutes.

**Figure 5.**
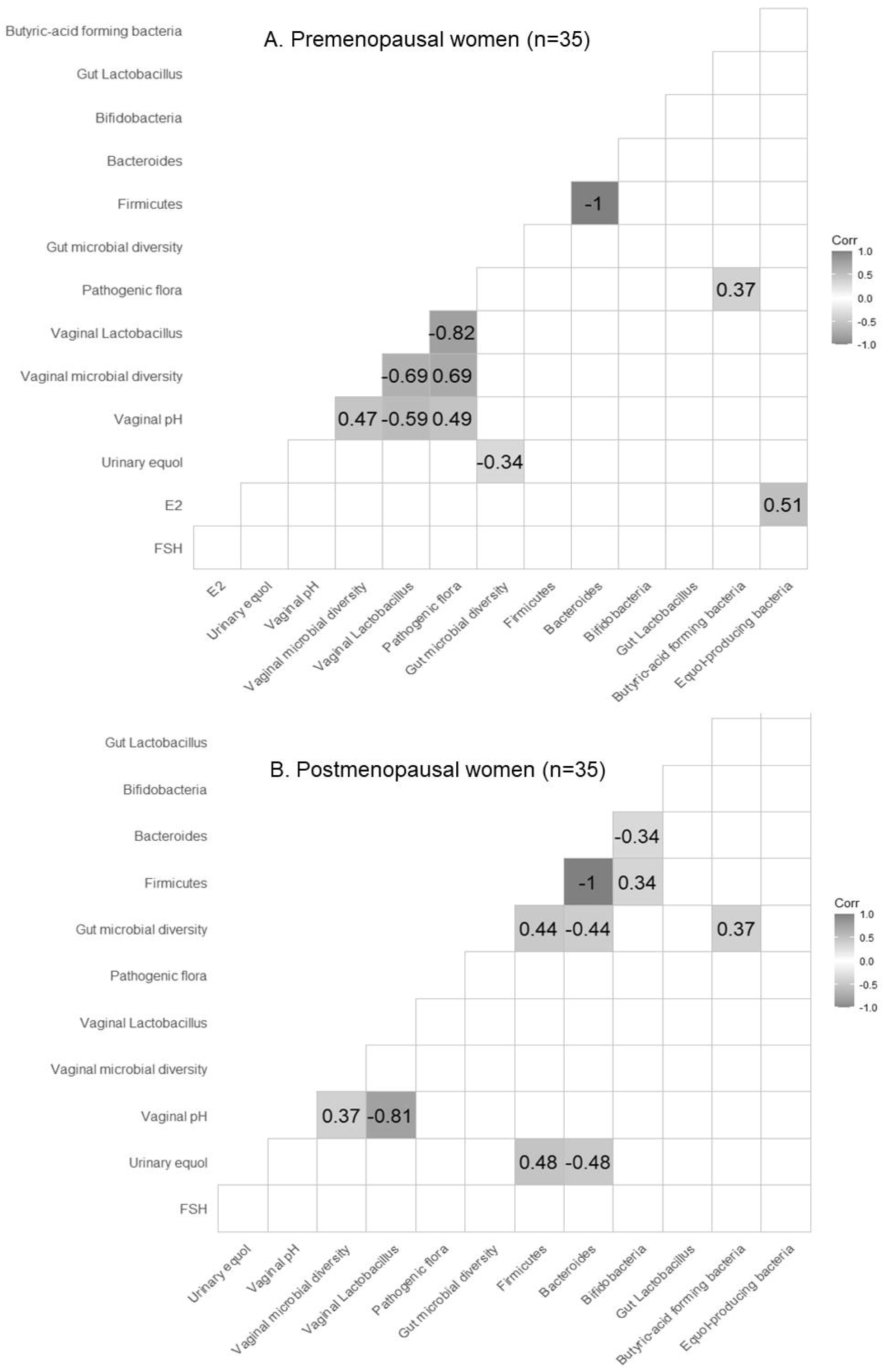
Factors affecting the vaginal microbiome. Only significant correlations (p<0.05) were described in the figure. Equol-producing bacterial populations and estrogen showed a positive association, while the relationship of equol and gut microbial diversity was negative in premenopausal women. Pathogenic flora had a strongly negative association with the vaginal *Lactobacillus* population in premenopausal women, but that association was not significant in postmenopausal women.

## Discussion

The ideal vaginal environment requires *Lactobacillus* as the dominant inhabitant and is under the control of blood estrogen levels. There were five community state types from CST I to CST V, depending on the composition of *Lactobacillus* species and their balance with the anaerobes in the vagina. CST IV was the type with the least lactobacillus composition or high diversity type, where the vaginal pH was at least 5.3. The CST type can vary with race, and the most dominant type in the Asian population is CST III or *Lactobacillus iners*. In this study, we found the same finding that CST III was approximately 35.02% and the most abundant type, as in previous studies. CST types found in postmenopausal women were I and III. The *Lactobacillus* population was 71.6% in premenopausal women but 10.3% in postmenopausal women, indicating the effect of declining estrogen on the *Lactobacillus* population. However, the proportions of gut *Lactobacillus* between the pre- and postmenopausal women were not different. Therefore, estrogen might have little or no effect on gut macrobacteria. There were seven common *Lactobacilli* between the gut and vagina. CST types seen in the gut were *Lactobacillus iners* (CST III) and *Lactobacillus gasseri* (CST II), but *Lactobacillus crispatus* (CST I) was not included. The characteristic of *Lactobacillus iners* (CST III) is its coinhabitation with pathogenic microflora. As it is the only species that can produce inerolysin, it may survive in unfavorable conditions. ^12, 13^ Therefore, after menopause in an unfavorable vaginal environment, it can still survive, whereas *Lactobacillus crispatus* (CST I) cannot survive. However, only 9% of *Lactobacillus iners* species can produce H_2_O_2_, ^14^ and it might have lower protection than other *Lactobacillus* species, which can produce both H_2_O_2_ and lactic acid to maintain the vaginal pH. In premenopsual women, there was competition between *L. iners* and *L. crispatus* inside the vagina, which was consistent with previous studies. ^15^ There were also relationships between common vaginal and gut *Lactobacillus* species. *L. rogosae*, which exists in the gut, also showed a relationship with vaginal *Lactobacillus*. Therefore, there seems to be crosstalk among the *Lactobacillus* species between the gut and the vagina.

The *Lactobacillus* population decreases with the decline in estrogen as women age. In this study, *L. iners* was still present even after menopause. It is thought to be well adapted to changes in the hormonal level. ^13^ In other words, *L. iners* might not entirely be under the control of the female sex hormone estrogen. We postulated that there could be crosstalk between *L. iners* in the gut and *Lactobacillus* species in the vagina after menopause. Regarding common bacteria in the gut and vagina that can cause bacterial vaginosis, there were positive relationships between them in both pre- and postmenopausal women. However, it was more prominent in postmenopausal women. Therefore, dietary habits and measures to improve the gut environment by increasing the *Lactobacillus* population in the gut might increase the population of *Lactobacillus* and reduce that of the bacterial flora in the vagina.

An interesting finding in this study was the relationship of equol to other factors. Equol, an isoflavone derivative whose chemical structure resembles estrogen, is produced from one type of isoflavone called daidzein. The benefits of equol range from relieved climacteric symptoms ^16, 17^ to prevention of bone density loss ^18^ and reduced risk of lifestyle-related diseases and cancers. ^19-23^ In our previous study with postmenopausal women, there was a strong relationship between urinary equol (equol producing ability) and gut microbial diversity. However, in this study, among premenopausal women, there was a positive relationship between estrogen and equol-producing bacteria, whereas the blood equol level had a negative relationship with gut microbial diversity.

We found similar findings to our previous studies: there was a positive correlation of urinary equol concentration with Firmicutes and a negative correlation with Bacteroides, and there was a positive relationship between Firmicutes and gut microbial diversity in postmenopausal women. However, these findings were not significant in premenopausal women. Therefore, there is a possibility that intrinsic equol production might be more interrelated with estrogen levels than gut microbiota in premenopausal women.

Between the gut and vaginal *Lactobacillus, L. gasseri* (CST II) showed a significant relationship in premenopausal women; however, it was *L. iners* in postmenopausal women. As there were differences in the *Lactobacillus* species or relationships with the gut and vagina due to age differences, the exact mechanism of the effect of intrinsic estrogen on the gut and vaginal microbiota requires further investigation.

The following are the limitations of this study. First, as it is a cross-sectional study, we could not determine the direction of relationships among the parameters in this study. However, this study highlighted the potential crosstalk between the vaginal and gut microbiomes and age-related or hormonal-dependent pathways in the composition of vaginal microflora. On the other hand, this is the first study that examined age- and hormonal-related differences in the composition of the gut and vaginal microbiome in both pre- and postmenopausal women.

## Conclusion

In postmenopausal women, a reduction in the *Lactobacillus* population and an increase in bacteria responsible for bacterial vaginosis were noted in the vaginal microbiota. However, there were no age-related or estrogen-related differences in the composition of gut microbiota. There was a relationship between the gut and vaginal microbiota in the populations of *Lactobacillus* and pathogenic flora, especially in postmenopausal women. Therefore, there could be crosstalk between the gut and vaginal microbiota. Improving the gut environment by dietary habits and other measures might also increase *Lactobacillus* and reduce the pathogenic microbiota in the vagina.

## Data Availability

All data produced in the present study are available upon reasonable request to the authors.

## Acknowledgments

We would like to acknowledge all the women who willingly participated in the study.

## Notes

Conflicts of Interest and Source of Funding: The Hamasite Clinic and Tokyo Midtown Medical Center receive administrative support from Advanced Medical Care Inc. Advanced Medical Care Inc. provided financial support for this research.

### Competing Interest Statement

The Hamasite Clinic and Tokyo Midtown Medical Center receive administrative support from Advanced Medical Care Inc. Advanced Medical Care Inc. provided financial support for this research.

### Clinical Protocols

https://upload.umin.ac.jp/cgi-open-bin/ctr_e/ctr_view.cgi?recptno=R000050146

### Funding Statement

This study was funded by Advanced Medical Care Inc.

### Author Declarations

The Institutional Review Board of Medical Corporation Shinkokai gave ethical approval for this work.

